# The Crucial Role of Predictive Models in Childhood Asthma care: Improving Outcomes Through Data-Driven Insights

**DOI:** 10.1101/2025.07.23.25332082

**Authors:** Aditya Chakrabarty, A K M Raquibul Bashar

## Abstract

**Background:** Asthma is one of the most prominent chronic diseases in children and one of the most challenging ailments to diagnose in infants and preschoolers in the United States. Predictive models can be instrumental in offering a data-driven approach to improve early diagnosis, personalize treatment strategies, and disease progression. By utilizing nationalized data, this study focuses on building and comparing high-performing analytical predictive models based on the 28 associated risk factors and identifying the most contributing factors influencing childhood asthma.

**Method:** Data came from the BRFSS (2011-2020) Asthma Call Back Survey (ACBS). The cross-sectional study included 9813 participants with a response rate of 65% (current asthma status positive). Respondents were randomly divided into training and testing samples. The grid-search mechanism was implemented to compute the optimum values of the hyper-parameters of the analytical eXtreme Gradient Boosting (XGBoost) model. The fitted XGBoost model was compared with four competing ML models, including support vector machine (SVM), random forest, LASSO regression, and GBM. The performance of all the models was compared using accuracy, AUC, precision, and recall. Variable importance plot (VIP) was used to measure the percentage of contribution of the predictors to the response, and Shapley Additive exPlanations (SHAP) plot was used to understand how the predictors are related to the outcome. Chi-square test was used to measure the association between the predictors and the outcome.

**Results:** Asthma diagnosis was found to vary by age group, with the highest prevalence in kindergarten age (31.44%). Of the five predictive models, the XGBoost was found to be the best performing model with AUC: 0.95, followed by random forest (AUC: 0.9345), GBM (AUC: 0.9341), SVM (AUC 0.9304), and LASSO (AUC 0.88); however, the random forest model was found to have the highest sensitivity (0.9786), and hence preferred for initial screening of asthma. The top two contributing predictors were overnight hospitalization visits and time since the last asthma medication, accounting for 24.62% and 20.92%, respectively, to the asthma status, from the VIP.

**Conclusion:** The analytical methodology of the model development was found to be instrumental in the discovery of behavior health-risk knowledge and to visualize the significance of predictive modeling from a multidimensional behavioral health survey. These insights can be instrumental in predicting different types of chronic lung diseases affecting people of all ages and can be useful for clinicians to diagnose asthma at an early stage, allowing for early intervention and proactive management.

## 1 Introduction

Asthma is one of the most common major non-communicable diseases and continues to impact the quality of life. Globally, asthma is ranked 16th among the leading causes of years lived with disability and 28th among the leading causes of burden of disease, as measured by disability-adjusted life years. Around 300 million people have asthma worldwide, and it is likely that by 2025, a further 100 million may be affected with this disease [29]. Despite progress in understanding asthma’s pathophysiology, the condition continues to pose a major public health challenge, with significant implications for individual and societal welfare [23]. Asthma is the most prominent chronic disease in children and one of the most challenging ailments to diagnose in infants and preschoolers. Castro-Rodriguez, Jose A., explains the usefulness of the Asthma Predictive Index (API) to address the recurrent wheezing of preschoolers/infants at the school-age [16]. There are multiple risk factors (deficiency in vitamin D, tobacco smoke exposure, air pollution, maternal anxiety or depression, allergen exposure, genetic risk factors, etc.) that have been found to be associated with asthma, and the differences in distributions of these risk factors can be attributed to the differences in prevalence that has a declining pattern in some parts of the world [54]. Although asthma is often manageable through inhaled medications, its clinical course varies, with some individuals experiencing persistent or recurrent symptoms into adulthood [40]. Consequently, identifying predictive factors and constructing models to forecast asthma development in children is crucial, potentially facilitating early intervention and personalized management strategies.

Machine learning techniques have become promising tools for predicting and managing various health conditions, including asthma, owing to their capacity to analyze complex datasets and discern intricate patterns not readily apparent via traditional statistical methods [31][21]. Leveraging large datasets, machine learning algorithms can identify risk factors, predict disease onset, and personalize treatment approaches, potentially improving patient outcomes and reducing healthcare costs [12]. The Behavioral Risk Factor Surveillance System serves as a valuable data source for studying health-related behaviors and risk factors among the US population [26]. Applying machine learning techniques to BRFSS data can yield valuable insights into predicting childhood asthma, potentially leading to more effective prevention and management strategies [14]. In recent years, researchers are prone to using sophisticated data-driven machine learning and decision-making models in applied sciences because of their high predictive power and learning abilities from data [11], [22], [19], [55], [20], [18], [53], [17], [7], [5, 8], [6]. [21] developed a predictive model for pancreatic cancer data using the XGBoost algorithm (eXtreme Gradient Boosting) and validated the model’s performance by comparing it with ten deep neural network (DNN) models [50], [24] grown sequentially with different activation functions and optimizers [49], [1]. Daines et al. studied seven existing prediction models to support the diagnosis of asthma in children and adults in primary care and found that patients who reported wheezing, variability of symptoms, and a history of allergy or allergic rhinitis were associated with asthma [28]. Colicino, Silva, et al. used validated tools for predicting childhood asthma based on MEDLINE and EMBASE (1946-2017) data for all available childhood asthma prediction models, and recommended the inclusion of larger cohorts, and more predictors into the model, along with more advanced statistical techniques to obtain better predictive accuracy [27].

## 2 Materials and Methods

### 2.1 Data

Data used in this study were acquired from the CDC (Centers for Disease Control and Prevention) website https://www.cdc.gov/brfss/acbs/index.htm. For the purpose of the study, we visited the survey data and documentation module of the BRFSS database. Then we selected the Asthma Return Visit Survey (ACBS) sub-database and downloaded the 2011 through 2020 Child Return Visit Survey Data. The nature of the study design was cross-sectional. During the study, data were collected from 9,9813 children aged 1 to 18 years from the BRFSS Asthma Callback Survey (ACBS) system [62, 61]. Call-back survey data were collected on all states in the US and its territories. So, the developed predictive models can be generalized to the US children (Ages: 1-18) population. Since the data was publicly available with de-identified information, we weren’t required to gain any privacy consent.

### 2.2 Outcome Variable

The outcome variable for the study “current asthma status” (Label: *Still Have Asthma Combined BRFSS and Call Back*) was selected as the outcome variable to determine whether the children still had asthma during the time of the call-back interview. Cross-sectional data [60] were analyzed in 9813 participants, including 6442 (65.6%) with current asthma status positive, from the BRFSS ACBS system.

#### 2.2.1 Predictors

After acquiring data from the CDC website, 28 predictors were selected as the initial list of risk factors based on the data code-book and questionnaires available on the website, as mentioned earlier. The Least Absolute Shrinkage and Selection Operator (LASSO) regression [52] [33] was used to select the final list of variables/risk factors to be included in the model-building process. Based on this statistical variable selection technique, 25 variables were selected where all risk covariates were categorical except *Age*. This variable was treated as an ordinal variable with 5 categories. The predictor variables included in this study are listed in the table 1

**Table 1:**
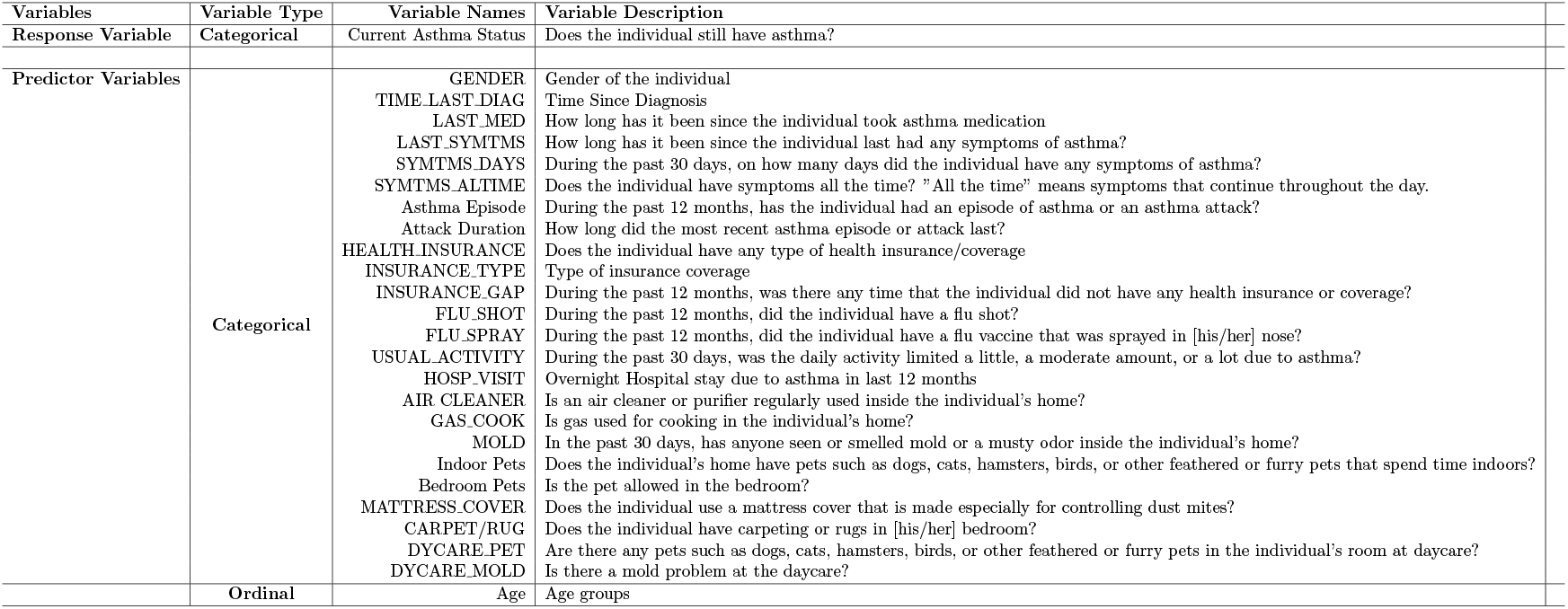
List of variables included in the Prediction Model Building Scheme.

### 2.3 Machine Learning Models

The machine learning models used in this study are briefly summarized as follows:

#### GBM

Gradient Boosting Machine (GBM) is a powerful machine learning technique that combines predictions from multiple decision trees to generate final predictions. This ensemble method is commonly used for both classification and regression problems, as it can effectively model complex, nonlinear relationships in data. GBMs work by iteratively building a sequence of decision trees, where each new tree focuses on correcting the errors of the previous one. This iterative process leads to an ensemble that is more accurate and robust than any single tree alone [46]. GBMs have been widely adopted across various industries and domains due to their ability to handle a wide range of data types and deliver strong predictive performance. The core principle behind GBMs is to construct new base learners that are highly correlated with the negative gradient of the loss function associated with the entire ensemble. This allows the algorithm to concentrate on areas where the existing ensemble performs poorly, gradually improving the overall predictive power. GBMs offer flexibility in terms of the loss function, as researchers can choose from a variety of loss functions or even implement their task-specific loss function, making the algorithm highly customizable to any particular data-driven task [35] [37].

#### XGBOOST

One of the most popular and widely used GBM algorithms is extreme gradient boosting (XGBoost). In the field of machine learning, one algorithm that has attracted considerable attention is XGBoost, a highly efficient and scalable implementation of the gradient-boosting technique. XGBoost has been widely acclaimed for its outstanding performance in numerous machine learning and data mining competitions, securing its position as the go-to choice for ensemble methods [21] [43] [10]. Furthermore, the thorough examination of the parameter tuning process in XGBoost can be invaluable to researchers across different fields. The parameter tuning process can offer insights into how various parameter configurations impact the algorithm’s performance, empowering researchers to make well-informed choices when optimizing XGBoost for their specific data and modeling needs [44]. By gaining a deeper understanding of the nuances in parameter setting, researchers can harness XGBoost’s flexibility to achieve exceptional predictive accuracy and efficiency within their respective domains.

#### LASSO

The Least Absolute Shrinkage and Selection Operator (LASSO) is a statistical technique primarily used for feature selection and data model regularization. In contrast to traditional regression models, LASSO has the unique capability to shrink certain regression coefficients to zero, effectively performing feature selection [57] [59]. This characteristic makes LASSO a valuable tool for predictive modeling, as it enables researchers to identify the most influential variables that drive the prediction of the target variable. The academic literature has extensively documented the utility of the LASSO algorithm in predictive modeling [39] [2]. LASSO proves particularly advantageous when working with high-dimensional datasets, where the number of predictor variables exceeds the number of observations. In such scenarios, the LASSO method can help prevent overfitting by selecting only the most relevant features, leading to more robust and generalizable predictive models. This feature selection ability makes LASSO a valuable tool for developing accurate and parsimonious predictive models, especially in fields with complex, high-dimensional data. Furthermore, multicollinearity, a prevalent issue in regression analysis where predictor variables are highly correlated, can be addressed with LASSO. LASSO can successfully reduce the impacts of multicollinearity by reducing some regression coefficients to zero, producing models that are more stable and comprehensible.

#### Random Forest

The random forest (RF) technique is a powerful and widespread machine learning approach that combines the outputs of multiple decision trees to generate a single predictive outcome. This ensemble method is particularly advantageous for both classification and regression tasks, as the aggregation of multiple models enhances the overall accuracy and robustness of the predictions [13]. The algorithm operates by creating a collection of individual decision trees, each trained on a random subset of the training data and a random subset of the features. The final prediction is then derived by aggregating the outputs of all the individual trees, either through majority vote or averaging. This ensemble strategy helps to mitigate the risk of overfitting and provides a more stable and reliable model compared to a single decision tree [58]. One of the key strengths of the random forest algorithm is its ability to handle large datasets with a high number of features [4]. Unlike traditional regression methods, the random forest algorithm can automatically detect and handle nonlinear relationships in the data, making it well-suited for complex and real-world problems. Additionally, the algorithm can accommodate a variety of data types, including numerical, categorical, and textual data, and is robust to outliers and missing values [3]. Furthermore, the random forest algorithm provides a measure of variable importance, which can be useful for feature selection and understanding the relative contribution of different variables to the model’s predictions [34]. This information can assist researchers and practitioners in identifying the most crucial factors driving the model’s outputs, which can be valuable for both interpretability and model optimization.

#### SVM

Support Vector Machines (SVM) are a powerful supervised machine learning algorithm widely used for classification and regression tasks [51] [15]. These models aim to identify an optimal hyperplane that maximally separates data points belonging to different classes, effectively acting as a decision boundary for classifying new, unseen instances. Furthermore, support vector machines are renowned for their computational efficiency, as the optimization problem they seek to solve is convex in nature and can be addressed using well-established algorithmic techniques [47]. A key advantage of SVMs is their ability to generalize well, even with relatively small training datasets, making them particularly useful for domains where data collection is challenging or expensive. Additionally, SVMs have demonstrated effectiveness in handling high-dimensional data and complex non-linear relationships by leveraging kernel functions, which transform the input data into a higher-dimensional space where linear separation becomes feasible [51] [36].

### 2.4 Model building, validation, and evaluation

To develop and evaluate the predictive models, the dataset was randomly partitioned into a training set (60%) and a test set (40%). All stages of the model development were performed independently for the predictive algorithms described under section 2.3. Model development and tuning were performed exclusively on the training dataset to avoid overfitting and enhance the performance of the models. To optimize the performance of each algorithm, a grid search with cross-validation was employed using the training dataset [21] [9]. For the LASSO model, regularization techniques were implemented to improve feature selection and mitigate overfitting. The ensemble methods (XGBoost, RF, and GBM) were fine-tuned to achieve a balance between model complexity and generalization capability. For the Support Vector Machine model, various kernel functions were tested to maximize classification accuracy. The predictive performance of each trained model was evaluated using an independent test set. Four key metrics were employed: sensitivity, specificity, area under the receiver operating characteristic (ROC) curve, and overall accuracy. Sensitivity and specificity were used to evaluate the model’s ability to accurately classify positive and negative instances, respectively. The AUC was used to determine the model’s discriminative capacity across various thresholds, while accuracy was used to summarize the proportion of correctly classified instances within the test dataset. By evaluating these diverse metrics, a comprehensive assessment of model performance was warranted, particularly in balancing the trade-offs between false positives and false negatives. The model that produced the highest AUC and balanced sensitivity and specificity was considered the most effective for prediction in this study.

## 3 Results

In this section we describe different performance matrices based on the five machine learning (ML) models. Table 2 summarizes the performance of five machine learning models—XGBOOST, LASSO, SVM, Random Forest (RF), and Gradient Boosting Machine (GBM), in predicting childhood asthma, evaluated using four metrics: accuracy, specificity, sensitivity, and area under the curve (AUC). As the table illustrates, XGBOOST demonstrated the best overall predictive accuracy (90.67%, AUC: 94.85%) indicating strong discriminative ability and reliable identification of true asthma cases, followed by LASSO (90.59%), SVM (90.34%), RF (90.16%), and GBM (88.99%). Also, XGBOOST has the highest AUC value followed by RF, GBM, SVM, and LASSO. The prediction accuracy and performance were found to be high, consistent, and robust for all five models (Table 2, and Figure 1). The random forest model was found to have the highest sensitivity (97.86%) followed by the XGBOOST (96.55%), and LASSO (96.47%). GBM provided a balanced trade-off between sensitivity (93.21%), specificity (80.93%), and AUC (93.41%), making it a reliable alternative.

**Table 2:**
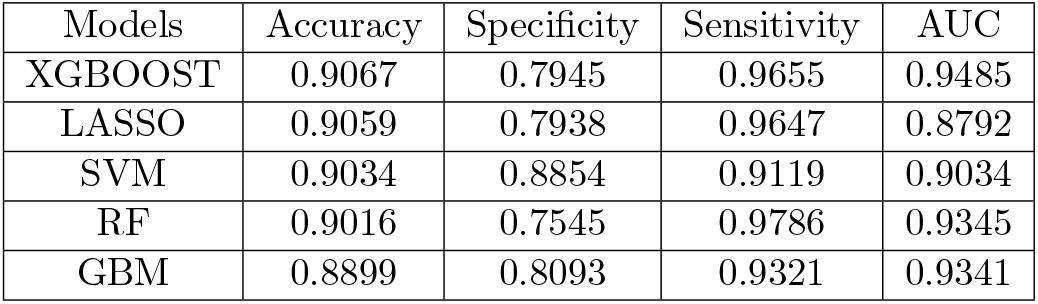
Comparing Different ML Models via Four Performance Matrices.

| Models | Accuracy | Specificity | Sensitivity | AUC |
| --- | --- | --- | --- | --- |
| XGBOOST | 0.9067 | 0.7945 | 0.9655 | 0.9485 |
| LASSO | 0.9059 | 0.7938 | 0.9647 | 0.8792 |
| SVM | 0.9034 | 0.8854 | 0.9119 | 0.9034 |
| RF | 0.9016 | 0.7545 | 0.9786 | 0.9345 |
| GBM | 0.8899 | 0.8093 | 0.9321 | 0.9341 |

**Figure 1:**
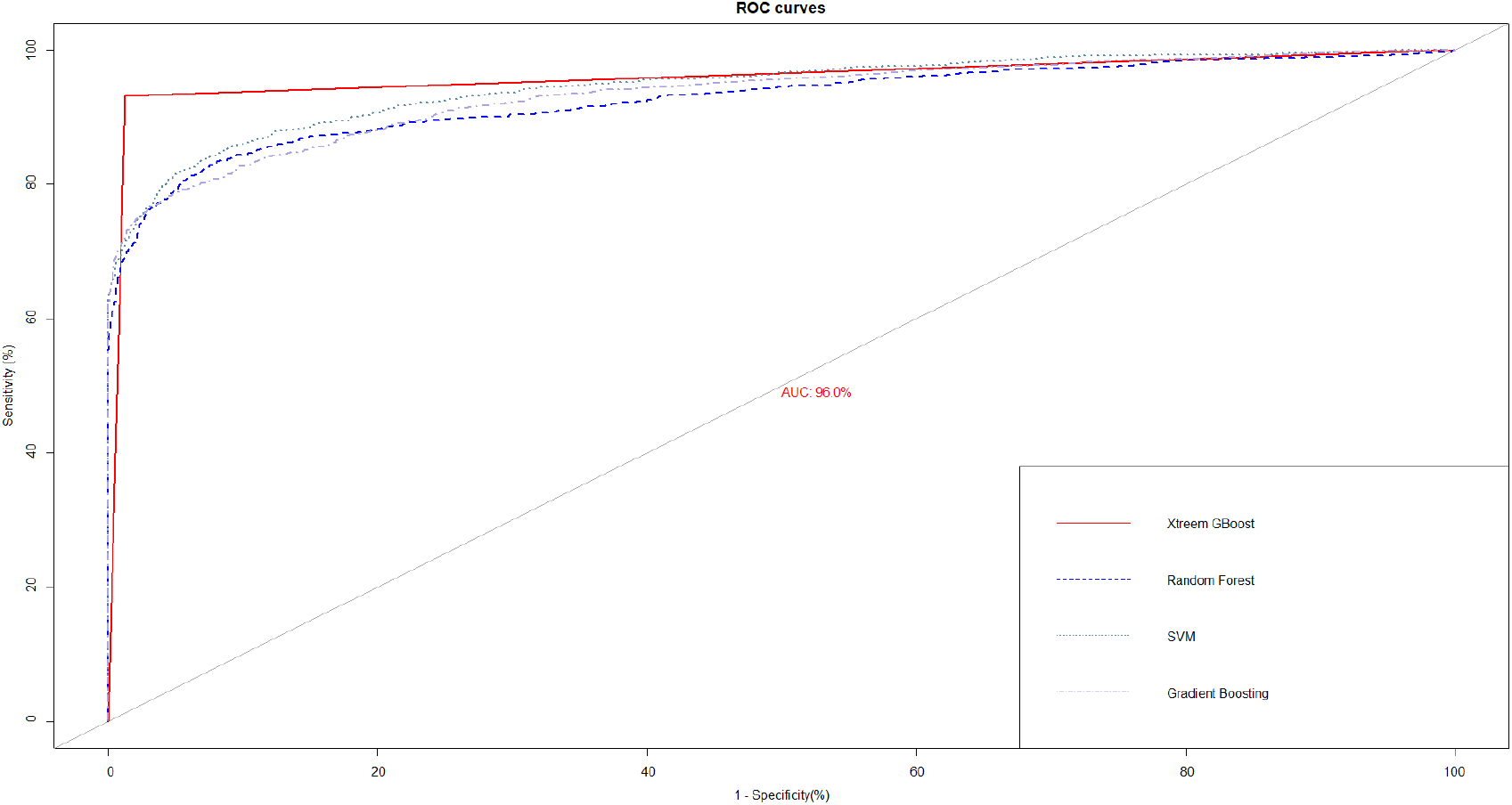
Model Comparison via ROC Curves.

The values of the comparison matrix are presented as shown in the following table (2)

Analyzing the main features influencing the performance of a predictive model is a critical component of statistical modeling. This task is especially vital in clinical settings, where comprehending the decision-making rationale of machine learning models is essential for making informed decisions [42] [56]. One approach to addressing this challenge is the use of SHAP (Shapley Additive exPlanations) values, which offer a comprehensive measure of feature importance [41]. Shapley values, which originate from game theory, provide a distinctive viewpoint on feature importance by evaluating the marginal contribution of each feature to the overall model performance [48]. This framework facilitates the identification of the key factors driving model predictions, enabling researchers and practitioners to gain a deeper understanding of the underlying mechanisms and potentially address issues such as overfitting or heterogeneity.

The SHAP plot in Figure 2 helps to explain the associations of the features with the asthma status in terms of SHAP values. In the SHAP plot, the SHAP values are used to analyze the strength of the association between individual feature contributions and model predictions. This helps to interpret how much a particular feature influences the model’s decision-making. A high SHAP value for a specific feature indicates a strong relationship with the model’s output, implying that changes in that feature can significantly impact model predictions. From the SHAP plot, we see that not overnight hospitalization (hospital visit No-Asthma) has a strong negative impact on the asthma status (reduces the likelihood of asthma prediction). When a patient has no asthma-related hospital visits, it strongly decreases the likelihood of an asthma diagnosis. The narrow violin plot suggests this effect is consistent. Time since the last asthma medication less than 1 day (LAST MED *<* 1 day) and time since the last asthma medication less than 3 months (LAST MED *<* (1-89 days)) have a notable positive impact on asthma prediction.

**Figure 2:**
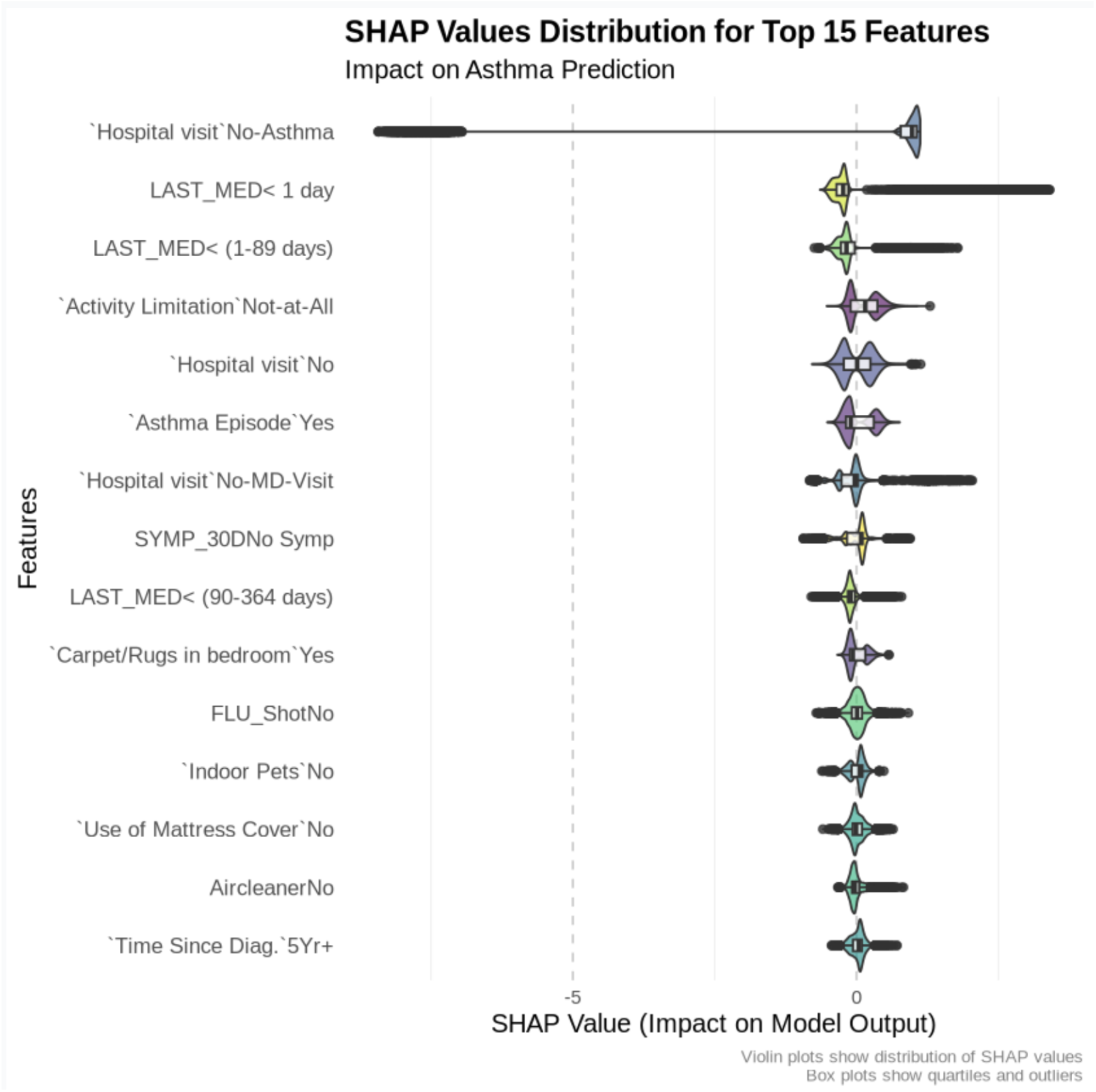
Shapley Plot for Feature Importance.

Activity limitation has a slightly negative impact on the outcome; patients reporting no activity limitations (Activity Limitation Not-at-All) reduced the predicted probability of asthma. No hospital visit in general has a negative impact. A recent asthma episode (Asthma Episode Yes) is strongly associated with asthma prediction. No MD visit (No-MD-Visit) has been found to have a dominating positive impact on the predicted outcome. The positive association suggests a higher risk of asthma-related complications when hospital visits do not involve an MD consultation. A negative association with the predicted asthma outcome was noted for the individuals with no asthma symptoms (SYMP 30D) in the last 30 days. Less recent medication use (LAST MED *<* (90-364 days)) was found to have a negative influence on asthma prediction, suggesting that medication use within the past 3 to 12 months might decrease the likelihood of asthma prediction. Some of the environmental factors (carpet/rugs in the bedroom, indoor pets, and air cleaner) were found to be significantly associated with asthma prediction. Not having the flu shot was found to be moderately associated with the predicted likelihood. Being diagnosed with asthma more than 5 years ago (time since the diagnosis 5 yr+) has a slightly negative association with the asthma prediction, which is evident from the distribution of the SHAP values (leans toward the left side of zero).

Table 3 shows the top fifteen contributing predictors arranged according to the importance score and % of contribution to childhood asthma that is also evident from Figure 3. The top four contributing predictors, according to the random forest (RF) algorithm are overnight hospitalization visits, time since the last asthma medication used, hindrance of usual activity due to asthma, and time since last asthma symptom, accounting for 24.62%, 20.92%, 12.19%, and 9.03% respectively, to the outcome.

**Table 3:** % Contribution of the Top Fifteen Predictors to the RF Model.

| Predictors | Importance | % Contribution |
| --- | --- | --- |
| Hospital Visit | 52.10 | 21.56 |
| LAST_MED | 44.24 | 18.32 |
| Activity Limitation | 25.78 | 10.67 |
| LAST_SYMTMS | 19.11 | 7.91 |
| Asthma Episode | 14.73 | 6.10 |
| Time Since Diag. | 12.51 | 5.18 |
| ATTACK/EPI.DUR | 11.38 | 4.71 |
| SYMP_30D | 11.22 | 4.64 |
| SYMTMS_ALTIME | 10.54 | 4.36 |
| DYCARE.MOLD | 9.90 | 4.10 |
| Age | 9.82 | 4.07 |
| DYCARE.PET | 8.70 | 3.60 |
| INSURANCE.GAP | 4.57 | 1.90 |
| Indoor Pets | 3.53 | 1.46 |
| Bedroom Pets | 3.40 | 1.41 |

**Figure 3:**
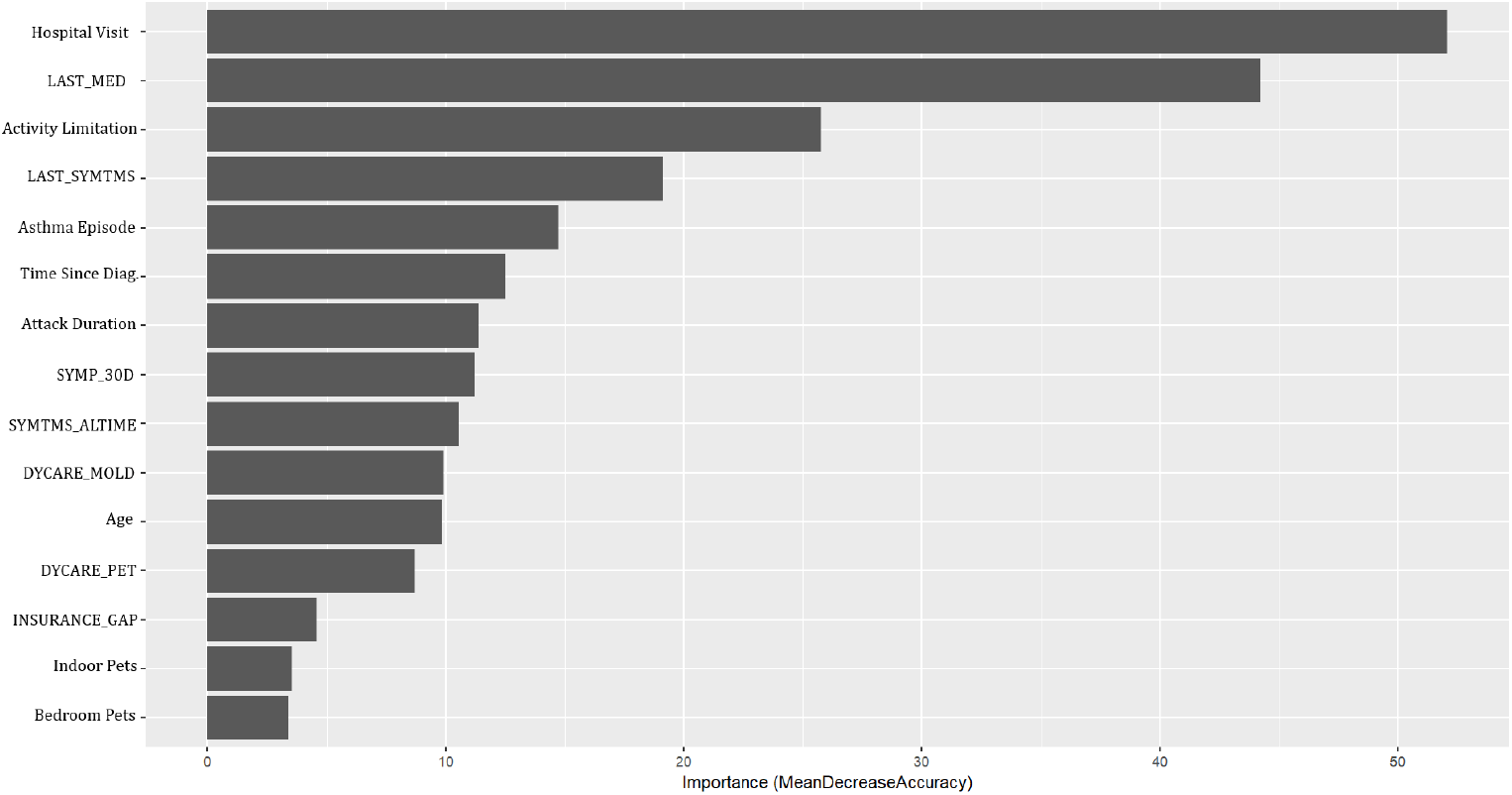
Top Fifteen Important Features Based on the Random Forest Model.

## 4 Discussion

Asthma is a chronic respiratory condition with significant public health implications due to its prevalence, associated healthcare costs, and impact on quality of life. In this study, five different predictive models were compared based on four different matrices. The XGBOOST emerged as the best-performing model overall, demonstrating the highest predictive accuracy (0.9067), sensitivity (0.9655), and AUC (0.9485) (Table 2). These metrics reflect the model’s strong ability to correctly classify both asthma and non-asthma cases, with particularly high effectiveness in detecting true positives. Where the accuracy can be misleading for unbalanced data, AUC deals with the class imbalance by evaluating the model’s ability to discriminate between classes across all possible thresholds [32]. Although the study data was not imbalanced (asthma yes = 65.65%, & asthma no = 34.35%), the AUC metric shows a consistent pattern regarding the models’ predictive power, and robustness to correctly discriminate the outcome categories.

In the context of childhood asthma prediction, high level of predictive performance is critical, as it ensures that most children at risk of developing asthma are accurately identified while minimizing misclassification. Accurate and early detection of children at risk of asthma can inform prompt interventions, such as environmental adjustments, early pharmacological intervention, and enhanced monitoring. The high AUC further validates the model’s capability to differentiate between affected and unaffected children, thereby supporting its application as a reliable tool for decision-making within pediatric primary care settings or public health screening initiatives.

The Random Forest (RF) model exhibited the greatest sensitivity among the competing models. High sensitivity is of extremely crucial in the early detection of asthma, as failure to identify affected individuals can lead to treatment delays, increased incidence of complications, and a worsen quality of life. This early diagnosis is vital for initiating early interventions, which can prevent disease progression and improve patient outcomes. High sensitivity also lessens the likelihood of false negatives by reducing misdiagnosis of patients. Missing an asthma diagnosis can lead to inadequate treatment, exacerbation, and potentially severe health consequences. The ensemble-based architecture of the RF algorithm facilitates the identification of nuanced patterns and interactions among various risk factors, thereby enhancing its ability to detect potential asthma cases, even in their developing stages or when symptoms are unclear, or ambiguous. However, the enhanced sensitivity of the RF model was associated with a associated reduction in specificity compared to the other models, potentially resulting in a higher rate of false-positive diagnoses. Despite this drawback, the prioritization of sensitivity is often warranted in the context of a screening instrument, particularly when applied to susceptible pediatric populations. Although initial over-identification may precipitate further evaluations, it ensures a minimal number of missed true cases, which constitutes an acceptable compromise given that early diagnosis can substantially improve long-term prognoses.

Important information about the factors affecting asthma diagnosis and severity was described using SHAP values. The model holds significant potential for informing public health interventions and policy-making due to its excellent predictive accuracy and interoperability. The SHAP analysis emphasizes how healthcare utilization, such as hospital stays and use of recent medication, are the significant predictors of asthma, reflecting the severity of the condition and management strategies. These results are consistent with previous research showing that frequent hospital stays and ER visits are indicators of poorly managed asthma [45] [30]. By incorporating the model into clinical decision-making processes, high-risk individuals may be identified early, allowing for rapid therapies to lessen the consequences associated with asthma. Asthma predictions are also found to be significantly influenced by environmental factors, including indoor dogs, carpets, and the absence of air purifiers, highlighting the importance of allergen exposure in asthma treatment. These findings highlight the necessity of environmental health initiatives aimed at lowering exposure to indoor allergens [25]. For example, especially in homes with asthmatic members, public health initiatives could encourage allergen-reducing practices including minimizing pet exposure, eliminating carpets, and utilizing HEPA filters [38].

Asthma predictive models have significant usefulness for several reasons. These predictive models can be useful in curating personalized treatment plans, and hence by improving the quality of life of the patients diagnosed with asthma. By predicting the course of asthma for individual patients, healthcare providers can tailor treatments to meet specific needs, improving outcomes and reducing unessential interventions, thus helping patients manage their condition better, leading to an enhanced quality of life. Effective asthma management guided by efficient predictive models can reduce hospital admissions and emergency visits, significantly lowering healthcare costs. Finally, these models can inform public health strategies and policies by identifying potential disease trends, contributing risk factors, and the impact of environmental or genetic factors on asthma prevalence and outcomes.

### 4.1 Limitation

One of the major limitations of this study is the lack of quantitative information available for building machine-learning models. Most of the variables in the survey questionnaire that we considered for model building were categorical in nature. While most ML algorithms perform better on quantitative features, including quantitative information in the development phase of the models might help in increasing the accuracy and relative performance. Also, our models were inherently limited by the features available in the BRFSS data. Because the BRFSS data is cross-sectional, causality couldn’t be determined between the contributing features and the outcome. Another constraint is that the self-reported BRFSS data may be influenced by social desirability and recall bias, potentially impacting the accuracy of our predictive models. Despite these limitations, a larger number of observations and Machine learning models such as XGBoost can potentially provide insight to comprehend the complex nonlinear relationship between potential attributes, and the targeted outcome.

## 5 Conclusion

The findings of the study suggests that machine learning models, specifically XGBOOST and random forest, hold considerable promise for improving early asthma diagnosis, facilitating prompt interventions, and enhancing health outcomes for children at risk. Future research should concentrate on validating these models in external cohorts, exploring integration into electronic health records, and assessing their real-world clinical utility. The incorporation of predictive algorithms into routine pediatric care has the potential to facilitate personalized risk assessment and reduce the burden of childhood asthma through proactive management.

## Data Availability

Data used in this study were acquired from the CDC (Centers for Disease Control and Prevention)

https://www.cdc.gov/brfss/acbs/index.htm

